# Polygenic heterogeneity across obsessive-compulsive disorder subgroups defined by a comorbid diagnosis

**DOI:** 10.1101/2021.05.21.21257530

**Authors:** Nora I. Strom, Jakob Grove, Sandra M. Meier, Marie Bækvad-Hansen, Judith Becker Nissen, Thomas Damm Als, Matthew Halvorsen, Merete Nordentoft, Preben B. Mortensen, David M. Hougaard, Thomas Werge, Ole Mors, Anders D. Børglum, James J. Crowley, Jonas Bybjerg-Grauholm, Manuel Mattheisen

## Abstract

Among patients with obsessive-compulsive disorder (OCD), 65-85% manifest another psychiatric disorder concomitantly or at some other time point during their life. OCD is highly heritable, as are many of its comorbidities. A possible genetic heterogeneity of OCD in relation to its comorbid conditions, however, has not yet been exhaustively explored. We used a framework of different approaches to study the genetic relationship of OCD with three commonly observed comorbidities, namely major depressive disorder (MDD), attention-deficit hyperactivity disorder (ADHD), and autism spectrum disorder (ASD). First, using publicly available summary statistics from large-scale genome-wide association studies, we compared genetic correlation patterns for OCD, MDD, ADHD, and ASD with 861 somatic and mental health phenotypes. Secondly, we examined how polygenic risk scores (PRS) of eight traits that showed heterogeneous correlation patterns with OCD, MDD, ADHD, and ASD partitioned across comorbid subgroups in OCD using independent unpublished data from the Lundbeck Foundation Initiative for Integrative Psychiatric Research (iPSYCH). The comorbid subgroups comprised of patients with only OCD (N = 366), OCD and MDD (N = 1052), OCD and ADHD (N = 443), OCD and ASD (N = 388), and OCD with more than 1 comorbidity (N = 429). We found that PRS of all traits but BMI were significantly associated with OCD across all subgroups (neuroticism: *p* = 1.19⨯10^*−*32^, bipolar disorder: *p* = 7.51⨯10^*−*8^, anorexia nervosa: *p* = 3.52⨯10^*−*20^, age at first birth: *p* = 9.38⨯10^*−*5^, educational attainment: *p* = 1.56⨯10^*−*4^, OCD: *p* = 1.87⨯10^*−*6^, insomnia: *p* = 2.61⨯10^*−*5^, BMI: *p* = 0.15). For age at first birth, educational attainment, and insomnia PRS estimates significantly differed across comorbid subgroups (*p* = 2.29⨯10^*−*4^, *p* = 1.63⨯10^*−*4^, and *p* = 0.045 respectively). Especially for anorexia nervosa, age at first birth, educational attainment, insomnia, and neuroticism the correlation patterns that emerged from genetic correlation analysis of OCD, MDD, ADHD, and ASD were mirrored in the PRS associations with the respective comorbid OCD groups. Dissecting the polygenic architecture, we found both quantitative and qualitative polygenic heterogeneity across OCD comorbid subgroups.

## 1 INTRODUCTION

Obsessive-Compulsive-Disorder (OCD) is a common, long-lasting and disabling neuropsychiatric disorder with an estimated lifetime prevalence of 1% to 3% (Weissman, 1998; U.S. International institutes of health (NIH), 2016). It is the fourth most common psychiatric disorder and has been ranked by the World Health Organization as being among the most disabling medical conditions world-wide as it can substantially impair the patient’s social, occupational and academic functioning (Murray et al., 1996). OCD is considered a complex disorder with its risk likely being influenced by hundreds to thousands of genetic variants scattered across the genome, with small to modest additive effects (Taylor, 2013; Craig, 2008). Genome-wide association studies (GWAS) in OCD have found suggestive evidence for some single nucleotide polymorphisms (SNPs) and genes that are potentially involved in its pathogenesis (International Obsessive Compulsive Disorder Foundation Genetics Collaborative (IOCDF-GC) and OCD Collaborative Genetics Association Studies and OCGAS, 2017). Yet, overall these findings remain rather inconclusive with no single genetic variant reliably replicating across individual studies (Sampaio et al., 2013; Bozorgmehr et al., 2017). These studies did, however, suggest that an increase in sample size will likely aid the identification of genome-wide significant loci, following the example of other psychiatric disorders like major depressive disorder (MDD) (Wray et al., 2018), attention-deficit hyperactivity-disorder (ADHD) (Demontis et al., 2019) or autism spectrum disorder (ASD) (Grove et al., 2019). Another reason for inconclusive findings may be that the majority of current studies of OCD do not account for or put enough emphasis on the heterogeneity of the disorder, though genetic findings may vary as a function of moderator variables (Kulminski et al., 2016; Mataix-Cols et al., 2005; Mattina and Steiner, 2016). One gene that is implicated in one subgroup of OCD patients may not be relevant for another, potentially making it more difficult to find true associations. As 65%-85% of OCD patients manifest another psychiatric disorder concomitantly or at some other time point during their lifetime (Nestadt et al., 2009; Tükel et al., 2002; Gillan et al., 2017), often presenting very different symptoms (Ortiz et al., 2016), it raises the question whether comorbid patients form distinct (genetic) subgroups. Nestadt et al. (2009) proposed a sub-classification of OCD based on comorbidity into three subgroups, with each group being associated with distinct clinical characteristics, prevalence rates, age-of-onsets, and sex-distributions. Dissecting OCD into more homogeneous and accurate sub-phenotypes based on comorbidity, may therefore lead to the successful identification of genetic risk variants for OCD (Kulminski et al., 2016; MacRae and Vasan, 2011).

In recent years, a variety of genetic studies have shown that OCD shares some genetic background with the neuropsychiatric disorders it co-occurs with (Cross-Disorder Group of the Psychiatric Genomics Consortium, 2013; O’Connell et al., 2018). The genetic correlation of OCD and tourette syndrome (TS) has been estimated at 0.41 (*SE* = 0.15) (Davis et al., 2013), with anorexia nervosa (AN) at 0.49 (*SE* = 0.13) (Yilmaz et al., 2020), with MDD at 0.21 (*SE* = 0.05) (Cross-Disorder Group of the Psychiatric Genomics Consortium, 2013), with ASD at 0.12 (*SE* = 0.08) (Cross-Disorder Group of the Psychiatric Genomics Consortium, 2013), and ranges between -0.17 (*SE* = 0.07) (Cross-Disorder Group of the Psychiatric Genomics Consortium, 2013) and 0.67 (*SE* = 0.09) (Goodman et al., 2020; Hirschtritt et al., 2018) for OCD and ADHD. With a quantitative genetic modelling approach Du Rietz et al. (2020) showed that the phenotypic association between ADHD and an externalising factor, also loading onto OCD, was largely influenced by genetics and it was demonstrated that both ADHD factors (inattentive and hyperactive/impulsive symptoms) were genetically related to OCD (Hirschtritt et al., 2018). PRS derived from ASD genetic data predicted 0.11% of the phenotypic variance in OCD (Guo et al., 2017). More recently, evidence for disorder-specific genetic associations has also been demonstrated. Peyrot and Price (2021) identified two SNPs distinguishing OCD and ADHD, and one SNP distinguishing OCD and ASD, using a newly developed method to quantify the genetic differences between psychiatric disorders by testing for differences in allele frequencies between cases of two disorders. It has also been shown that the majority of genes that have been implicated in OCD, ASD, schizophrenia (SCZ), and bipolar disorder (BP) are disorder-specific (O’Connell et al., 2018) and that the phenotypic differences between ADHD and OCD are reflected in altered DNA methylation at specific sites, pointing towards heterogeneous regulatory changes in both disorders (Goodman et al., 2020). As OCD shows such a high and specific genetic overlap with its comorbid neurodevelopmental and psychiatric disorders, while at the same time also presenting very unique genetic correlates, we explored whether OCD comorbid subgroups show a heterogeneous genetic architecture depending on the combination of co-occurring disorders.

In this paper we focused on the potential heterogeneity of OCD subgroups defined by comorbidity with MDD, ADHD, and/or ASD, as these disorders form the biggest comorbidity groups in the iPSYCH OCD sample. While MDD is the most commonly co-occurring diagnosis with OCD (approx. 15%-39.5% (Lochner et al., 2014)), ADHD occurs in approx. 6%-34% of OCD cases (Geller et al., 2004; Anholt et al., 2010) and OCD patients have a fourfold increased risk of developing ASD (Meier et al., 2015). Because specific markers associated with OCD have not yet been identified, we applied a variety of genome-wide analyses, neither looking for specific associated SNPs nor meta-analysing the iPSYCH samples with the current PGC OCD GWAS, as the sample-size increase would have only been marginal. Instead, in a first step we used publicly available summary statistics from the PGC to compare the genetic landscape of OCD patients to patients with either MDD, ADHD or ASD. We dissected similarities and differences in correlation patterns of the four disorders with 861 other phenotypes. In a second step we used an independent and previously unpublished OCD dataset from iPSYCH and compared the polygenic architecture of comorbid samples of patients with an OCD diagnosis and a further diagnosis of either MDD, ADHD, ASD or any combination thereof. We explored differences in polygenic risk score (PRS) load across the different OCD comorbid groups using a multivariate (multiple outcomes) multivariable (multiple covariates) regression, as introduced by Grove et al. (2019). As training datasets we used eight phenotypes from a variety of domains (*psychiatric, personality/psychological, anthropomorphic/metabolic, education*, and *other health risk*) that exhibited a range of differing correlation patterns with OCD, MDD, ADHD, and ASD. As OCD, MDD, ADHD, and ASD showed heterogeneous genetic patterns in the analyses in step one, we hypothesized that a) the comorbid OCD subgroups in the iPSYCH sample would show a heterogeneous association pattern with the PRSes, depending on the training dataset and the combination of comorbid disorders in the OCD subgroup, and b) that this heterogeneity would be in line with the correlation patterns between OCD, MDD, ADHD, and ASD and the PRS training phenotypes. We expected that the heterogeneity across OCD co-morbid subgroups in the PRS analysis would vary depending on whether the correlations of MDD, ADHD, and ASD showed the same or opposing directions as OCD with the traits used as a training dataset in the PRS analyses (see Figure 1 for an overview of performed analyses).

**Figure 1.**
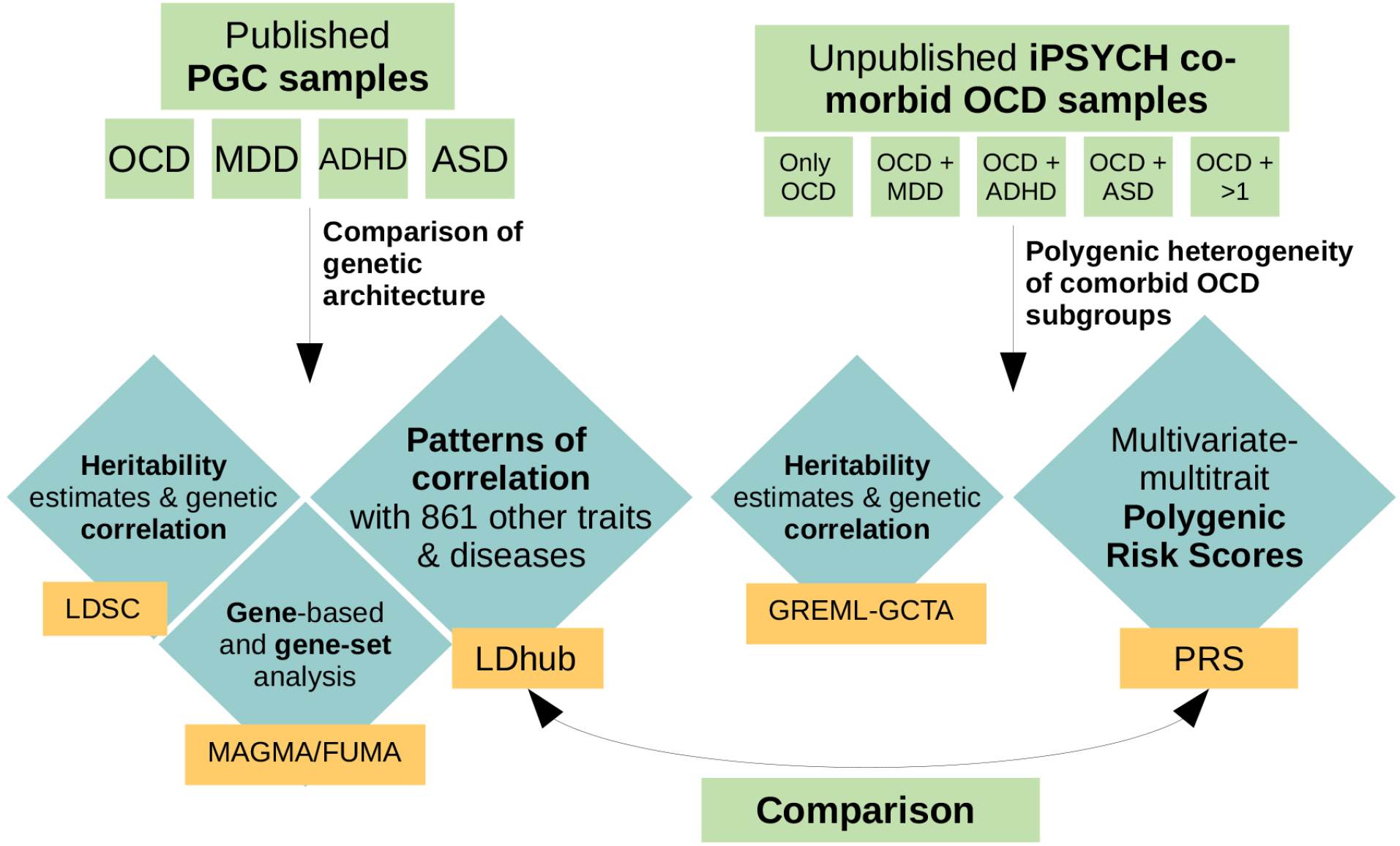
Schematic overview of performed analyses. For the first part of the analyses, we used previously published, publicly available GWAS summary statistics of obsessive-compulsive disorder (OCD), major depressive disorder (MDD), attention-deficit hyperactivity disorder (ADHD), and autism spectrum disorder (ASD) from the PGC. We used those datasets to compare the polygenic architecture of OCD, MDD, ADHD, and ASD by examining heritability estimates and genetic correlations (LDSC), their overlap in associated genes and gene-sets (MAGMA/FUMA), and compared each disorders genetic correlation pattern with 861 other traits and diseases (LDhub). For the second part of the analyses we used an independent and previously unpublished dataset from iPSYCH, comprising 2938 individuals with a diagnosis of OCD, of which 366 presented only a diagnosis of OCD, 1052 a diagnosis of OCD and MDD, 443 a diagnosis of OCD and ADHD, 388 a diagnosis of OCD and ASD, and 429 a diagnosis of OCD and more than one comorbidity. For these sub-groups we assessed heritability and genetic correlation estimates (GREML-GCTA) and examined the patterns of association of each comorbid OCD subgroup with eight different polygenic risk scores (PRS) based on a variety of phenotypes. In a last step we compared the patterns that evolved in the genetic correlation analysis in step one with the patterns of association that resulted from the PRS analyses of the OCD comorbid subgroups in step two.

## 2 METHODS

### 2.1 Subjects

#### 2.1.1 PGC samples

Publicly available European ancestry GWAS summary statistics of OCD, MDD, ADHD, and ASD were downloaded from the Psychiatric Genomics Consortium (PGC) website (see here). A description of sample sizes can be found in Table 1. Details about the cohorts and data processing have been described in the corresponding primary publications (OCD: International Obsessive Compulsive Disorder Foundation Genetics Collaborative (IOCDF-GC) and OCD Collaborative Genetics Association Studies and OCGAS (2017), MDD: Wray et al. (2018), ADHD: Demontis et al. (2019), ASD: Grove et al. (2019)).

**Table 1.** PGC & iPSYCH sample sizes, population prevalences and heritability estimates 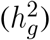. For PGC samples 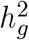 was estimated using LDSC, for the iPSYCH sub-samples univariate-GREML estimates of SNP-heritability are presented. All heritability estimates are reported on the liability scale (adjusted for population prevalence). Controls were the same for all iPSYCH subgroups. *Popprev*: population prevalence, *MC*: more than one comorbidity, *Ncont*.=Number of controls, *p*: p-value of 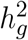. Within the MC group, 141 subjects are diagnosed with OCD, MDD, and ASD; 151 subjects are diagnosed with OCD, MDD, and ADHD, 153 subjects are diagnosed with OCD, ASD, and ADHD; and 34 subjects are diagnosed with OCD, MDD, ASD, and ADHD.

| Phenotype | Ncases | Ncontrols | Ntotal | Popprev. | $h_g^2$ (SE) | P |
| --- | --- | --- | --- | --- | --- | --- |
| **PGC** |  |  |  |  |  |  |
| OCD | 2688 | 7037 | 9725 | 0.03 | 0.29 (0.05) | - |
| MDD | 59851 | 113154 | 173005 | 0.15 | 0.09 (0.01) | - |
| ADHD | 19099 | 34194 | 53293 | 0.05 | 0.21 (0.01) | - |
| ASD | 18382 | 27969 | 46351 | 0.01 | 0.11 (0.01) | - |
| **iPSYCH** |  |  |  |  |  |  |
| onlyOCD | 366 | 10411 | 10819 | 0.01 | 0.29 (0.09) | 0.0003 |
| OCD+MDD | 1052 | 10411 | 11543 | 0.005 | 0.08 (0.03) | 0.0035 |
| OCD+ADHD | 443 | 10411 | 10901 | 0.002 | 0.04 (0.05) | 0.2312 |
| OCD+ASD | 388 | 10411 | 10840 | 0.0003 | 0.03 (0.04) | 0.2289 |
| MC | 429 | 10411 | 10890 | 0.0001 | 0.12 (0.03) | <0.0001 |

#### 2.1.2 iPSYCH comorbid OCD sample

In the scope of the *Danish OCD and Tourette Study* (DOTS) within *The Lundbeck Foundation Initiative for Integrative Psychiatric Research* (iPSYCH), Danish nation-wide population-based case-cohort samples were collected and genotyped. The study was approved by the Regional Scientific Ethics Committee in Denmark and the Danish Data Protection Agency. All analyses of the samples were performed on the secured national GenomeDK high performance-computing cluster in Denmark (see here) and Pedersen et al. (2018) for a detailed description of the overall cohort, array, genotyping, and quality control, here we give a brief summary. The iPSYCH sample comprised 2938 individuals with a diagnosis of OCD. All OCD patients that are included in the iPSYCH sample were either comorbid with one of the primary disorders in iPSYCH or were drawn from the population-based pool of controls. For each iPSYCH sample, DNA was obtained from the Danish Neonatal Screening Biobank (DNSB) at the Statens Serum Institut (SSI). Subsequent genotyping was performed in 23 batches on Illumina’s PsychChip v 1.0 array (Illumina, San Diego, CA, USA) at the Broad Institute of MIT and Harvard (Cambridge, MA, USA). Cases were identified amongst all individuals in iPSYCH (cases and controls) as individuals that met ICD10 diagnostic criteria for OCD (F42). Controls were randomly selected (for a 4 to 1 matching with cases) from the same cohort, and excluded individuals with a diagnosis of F42. Genotypes were processed using the *Rapid Imputation and COmputational PIpeLIne for Genome-Wide Association Studies* (ricopili) (Lam et al., 2020) performing stringent quality control of the data. Samples with call rates below 98% and individuals with a mismatch between sex obtained from genotyping and registered sex in the iPSYCH database were excluded. Related individuals were removed (randomly one individual per identified pair), principle component analyses were used to exclude ancestral outliers and the data was imputed using the 1000 Genomes Project phase 3 reference panel (The 1000 Genomes Project Consortium, 2015). The final dataset included 10411 controls and 2678 cases of which 366 were diagnosed with only OCD (*onlyOCD*), 1052 with OCD and MDD (*OCD+MDD*), 443 with OCD and ADHD (*OCD+ADHD*), 388 with OCD and ASD (*OCD+ASD*) and 429 with multiple comorbid disorders (*MC*) (see Table 1). Of the cases in the *MC* subgroup, 127 were diagnosed with OCD, MDD, and ASD; 140 with OCD, MDD, and ADHD; 129 with OCD, ASD, and ADHD; and 33 with OCD, ASD, ADHD, and MDD.

### 2.2 Statistical Analyses

#### 2.2.1 Gene-based and gene-set analysis

We performed gene-based- and gene-set association analysis of the PGC samples of OCD, MDD, ADHD, and ASD using the web-based tool *Functional Mapping and Annotation of Genome-Wide Association Studies* (FUMA) v1.3.1 (Watanabe et al., 2017) and *Multi-marker Analysis of GenoMic Annotation* (MAGMA) v1.6 (de Leeuw et al., 2015), employing a multiple regression model while accounting for linkage disequilibrium (LD) between the markers. For both analyses, the default MAGMA settings (SNP-wise model for gene analysis and competitive model for gene-set analysis) were applied. First, FUMA defines genomic risk loci on the basis of independent lead SNPs (with *r*^2^ *<* 0.1 between the independent lead SNPs), merging LD blocks that are physically closer than 250kb or overlapping into a single locus. Only SNPs in LD with a lead SNP and a minimum association p-value of 0.05 were included for further analysis. Each risk locus is represented by the top lead SNP with the minimum p-value in the locus. For MDD, ASD, and ADHD the minimum p-value of included lead SNPs was set to 5⨯10^*−*8^. Because the OCD GWAS had no SNPs exceeding the genome-wide threshold of 5×10^*−*8^ the threshold was arbitrarily lowered to 5⨯10^*−*6^. The minimum allele frequency (MAF) threshold was set to 0.01. 1000G phase 3 (The 1000 Genomes Project Consortium, 2015) was used as a reference panel to calculate LD across SNPs and genes and the MHC region was excluded. The gene-based p-values were computed by mapping SNPs to their corresponding gene(s) on the basis of their position in the genome. Positional mapping was based on ANNOVAR annotations and the maximum distance between SNPs and genes was set to 10kb. To correct for multiple testing, Bonferroni correction and false-discovery rate (FDR) were applied for gene-analysis and gene-set analysis, respectively. For OCD, input SNPs were mapped to 18709 protein coding genes, genome wide significance was defined at a Bonferroni corrected threshold of *p* = 2.67⨯10^*−*6^. FUMA tested curated gene-sets (c2.all) and gene ontology (GO) terms, using 10894 gene-sets for FUMA *≤* version 1.3.0 (ADHD) and 10655 gene-sets for FUMA *≥* version 1.3.1 (OCD, MDD, ASD). Gene-set p-values were computed using the gene-based p-values of all genes for each curated gene-set.

### 2.2.2 SNP-heritability estimates

SNP-heritability 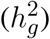 was estimated using LDSC (Zheng et al., 2017; Bulik-Sullivan et al., 2015b,a) for the PGC samples and univariate *genetic-relationship restricted maximum likelihood* (GREML) as implemented in *Genome-wide Complex Trait Analysis* (GCTA) (Yang et al., 2011; Lee et al., 2011) for the iPSYCH OCD subgroups, as sample sizes of the subgroups were too small for LDSC and raw genotype data was available. For LDSC, freely available precomputed LD scores based on the European ancestry samples of the 1000G phase 3 (The 1000 Genomes Project Consortium, 2015), restricted to HapMap3 SNPs, were used. Before the analysis, standard LDSC filtering was applied. Poorly imputed SNPs with *INFO < 0*.*9* were removed. For the conversion of observed-scale-to liability-scale estimates, previously reported disorder-specific prevalence rates were used (see Table 1).

For the comorbid iPSYCH samples the univariate GREML approach of GCTA was used. After removal of ancestry outliers, counts of each sub-phenotype were the following: *controls*: 10411, *onlyOCD*: 366, *OCD+MDD*: 1052, *OCD+ADHD*: 443, *OCD+ASD*: 388, *MC*: 429. A genetic relatedness matrix (GRM) was fitted, thereby providing relatedness estimates for all pairwise combinations of individuals. All indels were removed and the data was filtered on *genotype probability >* 0.8, *missing rate <* 0.01 and *MAF >* 0.05. GRM was estimated for each individual autosome and subsequently merged into a single GRM based on all autosomes. 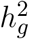 estimation for each OCD sub-phenotype was performed including the first four principle components (PCs) as continuous covariates together with any other PC that was nominally significantly associated to the phenotype. Waves were included as categorical indicator covariates. Lacking proper population prevalence estimates for subgroups, revalense rates for comorbid conditions were estimated by multiplying the prevalence for each comorbid disorder with the OCD prevalence (3%). The prevalence for the OCD subgroup with more than one comorbid disorder was estimated to be lower than any of the other prevalence rates at an arbitrary value of 0.01%, as the multiplication of more than two prevalence rates would strongly underestimate the true prevalence. Because at least one other psychiatric disorder is present in approximately two thirds of OCD patients (Tü kel et al., 2002; Gillan et al., 2017), the prevalence for *only OCD*, without any comorbid diagnosis, was set to 1% (one third of the general OCD prevalence). See Table 1 for a list of all population prevalence estimates.

#### 2.2.3 Genetic correlation estimates

Using LDSC (Bulik-Sullivan et al., 2015b,a) we estimated the genetic correlation (*r*_*G*_) of OCD with MDD, ADHD, and ASD. We further estimated each disorder’s genetic correlation with 861 other phenotypes using LDSC as implemented in LDhub (Zheng et al., 2017) (for 855 traits) and LDSC (for 6 additional traits not contained in the LDhub database). We then compared the correlation patterns that emerged for OCD to those of MDD, ADHD, and ASD.

Bi-variate GREML as implemented in GCTA was used to estimate the genetic correlation between the iPSYCH OCD subgroup samples. The controls were split proportionally in order to guarantee an independent control group for each comorbid subgroup in every pairwise comparison.

#### 2.2.4 Multivariate-Multitrait PRS analyses (PRS)

By applying multivariate (multiple outcomes) multivariable (multiple covariates) regression (Grove et al., 2019) we examined the distribution of PRSs based on OCD (International Obsessive Compulsive Disorder Foundation Genetics Collaborative (IOCDF-GC) and OCD Collaborative Genetics Association Studies and OCGAS, 2017), neuroticism (Nagel et al., 2018), anorexia nervosa (AN)(Watson et al., 2019), bipolar disorder (BP)(Stahl et al., 2019), Educational Attainment (EA) (Lee et al., 2018), body mass index (BMI) (Yengo et al., 2018), age at first birth (AFB) (Barban et al., 2016), and insomnia (Jansen et al., 2019), over the OCD comorbid subgroups. For the calculation of PRSs, the summary statistics of interest were clumped by applying standard ricopili parameters. Prior to clumping overlapping SNPs between the iPSYCH data and the external summary statistics were extracted and strand ambiguous A/T and C/G SNPs with a frequency between 0.4 and 0.6 were removed to avoid potential strand conflicts. PRS were generated at the default p-value thresholds of 5⨯10^*−*8^, 1⨯10^*−*6^, 1×10^*−*4^, 0.001, 0.01, 0.05, 0.1, 0.2, 0.5, and 1 as a weighted sum of the risk allele dosages. Prior to analysis scores were normalized. After the PRS were calculated, the scores were regressed onto the OCD subgroups to evaluate the genetic overlap between the phenotypes and the OCD subgroups. Batch effects from genotyping waves and PCs in the comorbid OCD data were adjusted for in the multivariate multivariable regression. The advantage of a multivariate regression is that it can handle a possible correlation among the PRSes, making it possible to test a great number of hypotheses across PRSes and subtypes. The approach is statistically very powerful which enables us to conduct these analyses even with sample sizes too small to conduct a GWAS or LDSC analysis.

## 3 RESULTS

### 3.1 Comparing the genetic architecture of OCD, MDD, ADHD, and ASD

#### 3.1.1 Gene & gene-set analysis

First, we performed gene-based- and gene-set association analysis of the PGC samples of OCD, MDD, ADHD, and ASD using MAGMA/FUMA, thereby looking for potential overlaps in associated genes and gene-sets between the four disorders. When looking at 13 genes that showed suggestive association for OCD (*p <* 1⨯10^*−*4^; strongest association for *KIT Proto-Oncogene Receptor Tyrosine Kinase* on chromosome 4, *p ≤* = 2.46⨯10^*−*7^) there was no evident overlap with significant genes of the other disorders (see Supplementary Table S1). Furthermore, no gene-set (*p* 9.7⨯10^*−*5^) overlapped between OCD, MDD, ADHD, and ASD (see Supplementary Table S2 for gene-set results of OCD).

#### 3.1.2 Heritability and genetic correlations

Next, we computed SNP heritabilities 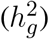 of OCD, MDD, ADHD, and ASD and calculated cross-trait genetic correlations (*r*_*G*_) between each pair of disorders using LDSC (Bulik-Sullivan et al., 2015b,a). OCD was significantly positively correlated with MDD (*r*_*G*_ = 0.23, *SE* = 0.07, *p* = 0.0005) and nominally significantly negatively correlated with ADHD (*r*_*G*_ = -0.17, *SE* = 0.07, *p* = 0.02), while the correlation between OCD and ASD did not reach significance (*r*_*G*_ = 0.12, *SE* = 0.08, *p* = 0.15). See Table 1 for all heritability and correlation estimates.

To investigate the extent of genetic overlap between OCD and an array of other phenotypes, we estimated its genetic correlations with 861 psychiatric and other medical diseases, disorders, and traits using bivariate LD score regression (Zheng et al., 2017; Bulik-Sullivan et al., 2015b,a). The same analysis was also performed for MDD, ADHD, and ASD as we were interested in similarities and differences in patterns of correlations between the four disorders. 777 (for OCD and ADHD), 778 (for ADHD), and 779 (for MDD) genetic correlations yielded interpretable results, the remaining estimations resulted in “NA”, due to small sample size and non-significant heritability. We therefore set the significance threshold to a Bonferroni-corrected p-value of 0.05/779=6.42⨯10^*−*5^. Of the tested diseases and traits, 49 were significantly correlated with OCD (to compare: 253 with MDD, 287 with ADHD, and 53 with ASD). 44 traits overlapped between OCD and MDD, 39 between OCD and ADHD, and 13 between OCD and ASD. Ten traits were significantly associated with all four disorders, of which six demonstrated the same direction of effect (see Supplementary Table S3).

A subset of the phenotypes that significantly correlated with OCD (except insomnia, which did not significantly correlate with OCD but with MDD and ADHD) was grouped into five categories: *psychiatric, personality/psychological, anthropomorphic/metabolic, education*, and *other health risk* (see Supplementary Figure S1-S5 and Supplementary Table S4). Across the four disorders (OCD, MDD, ADHD, and ASD), differences in their patterns of correlations emerged. While all four disorders generally showed positive associations with traits in *psychiatric disorders* and *personality/psychological* traits, ASD and ADHD exhibited fewer and/or lower significant associations than OCD and MDD (see Supplementary Figure S1 and S2). In the category of *anthropomorphic and metabolic* traits (see Supplementary Figure S3) OCD significantly correlated negatively with all reported parameters, while MDD correlated moderately positively and ADHD strongly positively with the same phenotypes. ASD did not significantly correlate with any of the listed traits. While OCD and ASD positively correlated with *education* traits (see Supplementary Figure S4), ADHD and MDD negatively correlated with all *education* parameters (for *no specific qualifications* the pattern was reversed). In the category of *other health risk* (see Supplementary Figure S5), OCD was almost exclusively negatively correlated, while MDD and ADHD were almost exclusively positively correlated with all phenotypes (for AFB the pattern was reversed).

From each category we selected one or two traits (see figure 2) to further evaluate how PRS based on a broad spectrum of phenotypes with varying patterns of correlations with OCD, MDD, ADHD, and ASD, partition across comorbid OCD subgroups (see below).

**Figure 2.**
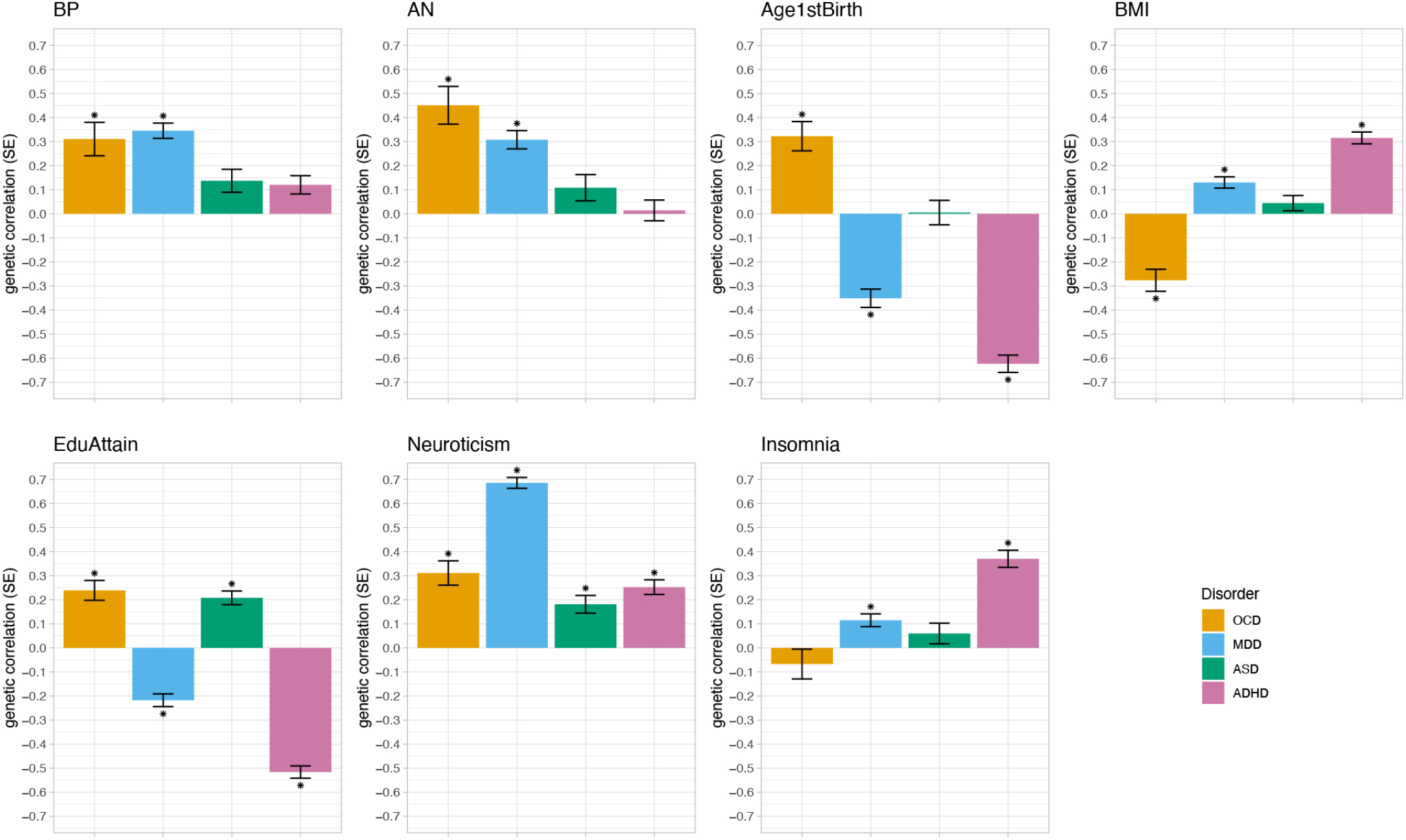
Differential genetic correlation patterns of OCD, MDD, ADHD, and ASD. Bivariate LD score regression was used for the analysis. * indicates statistical significance at a Bonferroni-corrected threshold of 6.42×10^*−*5^ corrected for 779 tests. The phenotypes correspond to the following data sets: bipolar disorder (BP) (Stahl et al., 2019), anorexia nervosa (AN) (Watson et al., 2019), age at first birth (AFB) (Barban et al., 2016), body mass index (BMI) (Yengo et al., 2018), educational attainment (EA) (Lee et al., 2018), neuroticism (Nagel et al., 2018), and insomnia (Jansen et al., 2019).

### 3.2 Dissection of the polygenic architecture of comorbid OCD subgroups

#### 3.2.1 Heritability and genetic correlations among the subgroups

Next, we explored the polygenic heterogeneity across OCD comorbid subgroups. We examined how 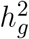 partitioned across the comorbid OCD subgroups and estimated the genetic correlation among these groups using GCTA (Yang et al., 2011). Univariate GREML analysis revealed significant 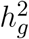 for the *onlyOCD, OCD+MDD*, and *MC* subgroups (see Table 1 for all 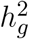 estimates). Pairwise comparisons of genetic correlations (*r*_*G*_) of the sub-phenotypes were estimated with bivariate GREML. Each subgroup demonstrated a high genetic correlation with all other subgroups (between 0.2 and 1; see Supplementary Table S5 for a list of the results). The *MC* subgroup had the lowest genetic correlations with all other subgroups, with *MC* and *OCD+MDD* demonstrating the lowest correlation between all pairwise-tests, while the *onlyOCD* group showed the highest correlations with all other comorbid groups. Standard errors were generally very high for all pairwise correlations.

#### 3.2.2 Cross-trait PRS analyses

To examine a possible polygenic heterogeneity of OCD, we further investigated how PRS trained on different phenotypes (*OCD, neuroticism, EA, AN, BP, BMI, AFB*, and insomnia) distribute across the iPSYCH OCD subgroups defined by a comorbid diagnosis of either MDD, ADHD, and/or ASD. Rather than selecting traits for the PRS analysis based on a theoretical background (e.g. clinical meaningfulness), they were chosen in view of their different correlation patterns with OCD, MDD, ASHD, and ASD. Thereby, we wanted to explore whether different correlation patterns with OCD, MDD, ADHD, and ASD would translate into differing patterns in the PRS analysis across the OCD comorbid subgroups. The traits used as training datasets in the PRS analysis either showed a) a significant correlation with OCD, MDD, ADHD, and ASD in either the same direction (BMI) or differing directions (EA); or b) a significant correlation with OCD and either one (BP) or two (AFB, BMI, AN) other disorders; or c) no significant correlation with OCD but a significant correlation with two other tested disorders (insomnia). Further, OCD itself was included as a training dataset for the PRS analysis. With this selection of phenotypes we aimed to explore whether a heterogeneous genetic correlation pattern between a phenotype and OCD, MDD, ADHD, and ASD translates into heterogeneous PRS loadings in the OCD comorbid subgroups.

The PRS analysis can be read as a linear regression with the beta value indicating the mean level of PRS relative to the controls, adjusted for the other variables and covariates (first four principle components and batches). First, for each phenotype, it was tested whether the betas of the PRS analyses were significantly different from zero across all OCD comorbid subgroups. *neuroticism, BP, AN, AFB, EA, OCD*, and insomnia showed significant associations with the iPSYCH OCD samples (*p* = 1.19⨯10^*−*32^, *p* = 7.51⨯10^*−*8^, *p* = 3.52⨯10^*−*20^, *p* = 9.38⨯10^*−*5^, *p* = 1.56⨯10^*−*4^, *p* = 1.87⨯10^*−*6^, *p* = 2.61⨯10^*−*5^, respectively; see Supplementary Table S6). Of the eight phenotypes tested (*neuroticism, BP, AN, AFB, EA, OCD, BMI*, and insomnia) for association with the OCD comorbid subgroups, AFB (*p* = 2.29⨯10^*−*4^), EA (*p* = 1.63⨯10^*−*4^), and insomnia (*p* = 0.045) showed a significant heterogeneity across OCD subgroups. BP and AN were positively associated with all OCD subhroups, while the other traits showed significant associations with some of the OCD comorbid subgroups, but not with all (see Figure 3 and Supplementary Table S7). For AFB the strongest, though non-significant, positive associations were with the *onlyOCD* group (*Beta* = 0.099, *CIl* = -0.004, *CIu* = 0.202, *p* = 0.059), followed by the *OCD+ASD* (*Beta* = 0.056, *CIl* = -0.039, *CIu* = 0.15, *p* = 0.247) group. The strongest negative association was with the *OCD+ADHD* group (*Beta* = -0.188, *CIl* = -0.288, *CIu* = -0.088, *p* = 2.29⨯10^*−*4^), followed by the, though non-significant, *MC* group (*Beta* = -0.08, *CIl* = -0.176, *CIu* = 0.015, *p* = 0.098), and *OCD+MDD* group (*Beta* = -0.067, *CIl* = -0.13, *CIu* = -0.004, *p* = 0.037) (see Figure 3 and Supplementary Table S7 and S8 for results of all tested phenotypes). For EA, there was a strong negative association with *OCD+ADHD* (*Beta* = -0.232, *CIl* = -0.333, *CIu* = -0.131, *p* = 6.36⨯10^*−*6^) and a trend for a positive association with *OCD+ASD* (*Beta* = 0.086, *CIl* = -0.009, *CIu* = 0.180, *p* = 0.077), while the other subgroups demonstrated scores around zero. For the PRS based on insomnia the strongest positive association was with the *OCD+ADHD* (*Beta* = 0.208, *CIl* = 0.107, *CIu* = 0.309, *p* = 5.62⨯10^*−*5^) group, followed by the *MC* (*Beta* = 0.133, *CIl* = 0.037, *CIu* = 0.229, *p* = 5.62⨯10^*−*3^) and the *OCD+MDD* group (*Beta* = 0.084, *CIl* = 0.021, *CIu* = 0.148, *p* = 9.39⨯10^*−*3^).

**Figure 3.**
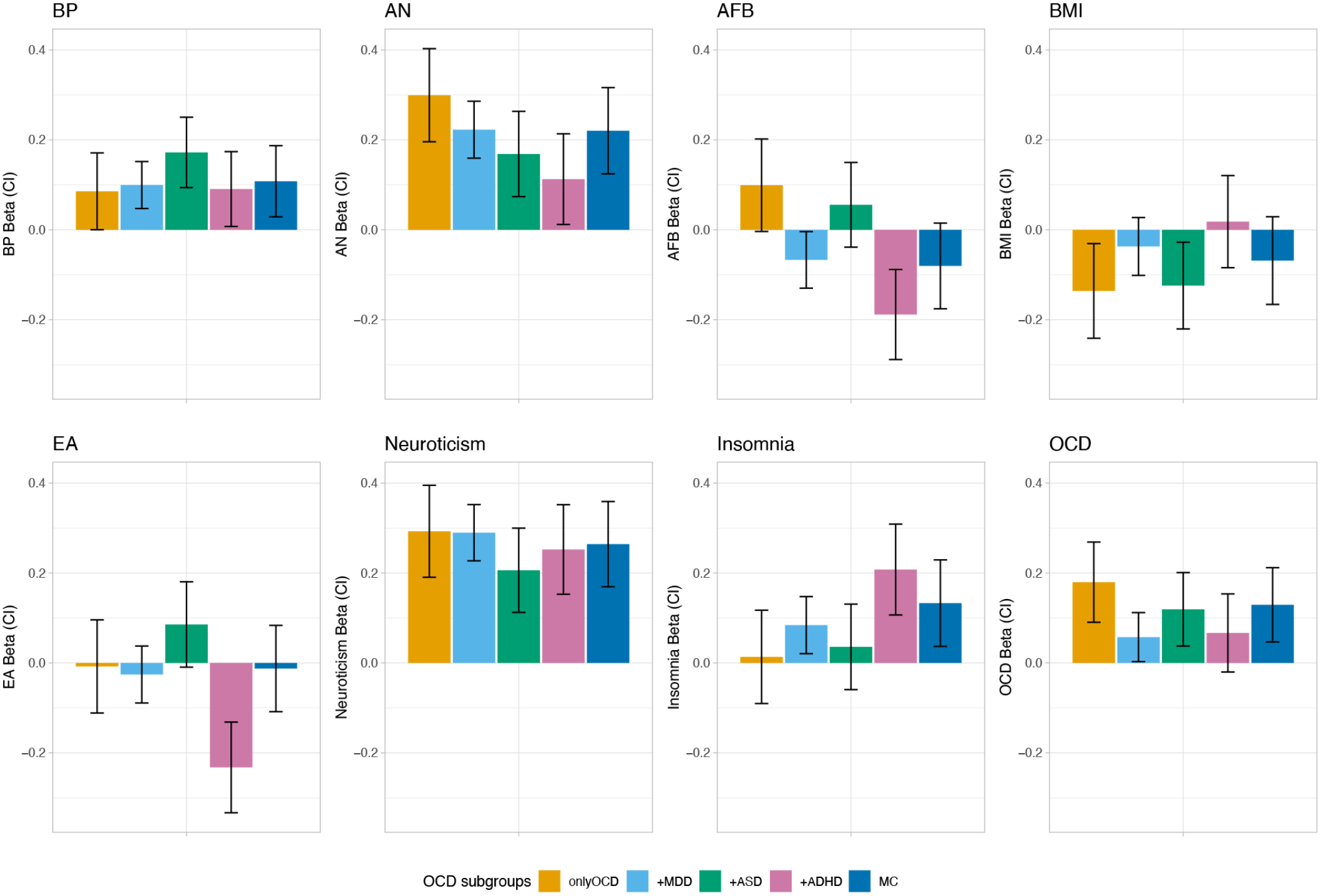
PRS profile across distinct comorbid OCD subgroups in the iPSYCH sample. The bars show coefficients from multivariate multivariable regression of the eight normalized PRS over OCD subgroups, adjusting for the first four PCs and for 23 waves, whiskers indicate 95% confidence intervals (CI). Results are shown for eight different phenotypes: bipolar disorder (BP) (Stahl et al., 2019), anorexia nervosa (AN) (Watson et al., 2019), age at first birth (AFB) (Barban et al., 2016), body mass index (BMI) (Yengo et al., 2018), educational attainment (EA) (Lee et al., 2018), neuroticism (Nagel et al., 2018), obsessive-compulsive disorder (OCD) (International Obsessive Compulsive Disorder Foundation Genetics Collaborative (IOCDF-GC) and OCD Collaborative Genetics Association Studies and OCGAS, 2017), and insomnia (Jansen et al., 2019).

## 4 DISCUSSION

In the present study we first looked at genetic similarities and differences between OCD and the three psychiatric disorders MDD, ADHD, and ASD, with a specific emphasis on the genetic correlation patterns of each of the four disorders with 861 somatic and mental health phenotypes. In a second step we used genome-wide data of an independent set of OCD patients from iPSYCH for which we defined five OCD subgroups based on the patients’ comorbidity with MDD, ADHD and/or ASD (*onlyOCD, OCD+MDD, OCD+ADHD, OCD+ASD*, and *MC*). Using eight different traits (BP, AN, AFB, BMI, EA, neuroticism, insomnia, and OCD) as training data sets, we applied PRS analysis across the comorbid OCD subgroups. We hypothesized that a) the comorbid OCD subgroups show a heterogeneous association pattern with the PRSes, depending on the training dataset and the combination of comorbid disorders in the OCD subgroup, and b) that this heterogeneity is in accordance with the correlation patterns between OCD, MDD, ADHD, and ASD and the PRS training phenotypes. More precisely, we expected that the heterogeneity across OCD co-morbid subgroups in the PRS analysis would vary depending on whether the correlations of MDD, ADHD, and ASD showed the same or opposing directions compared to OCD with the traits used as a training dataset in the PRS analyses (e.g. if MDD showed a positive correlation with trait A and OCD a negative correlation with trait A, we expected that the onlyOCD group would show a lower association with the PRS based on trait A than the comorbid subgroup of OCD+MDD, while we hypothesized that a positive correlation of both, MDD and OCD, with trait A would translate into either an increased or similar association of the PRS based on trait A with the OCD+MDD comorbid subgroup compared to the onlyOCD group).

The genetic correlation patterns that emerged in the first part of the analysis are generally in accordance with symptomatic and clinical observations of OCD, MDD, ADHD, and ASD patients. All four disorders displayed positive associations with most of the other psychiatric disorders and with personality/psychological parameters, such as BMI, *worry*, and *tense feelings*. The genetic correlation of OCD wanthropomorphic and metabolic traits was negative, while MDD and ADHD showed a positive correlation. This is in line with the observation that OCD is genetically positively correlated with AN (The Brainstorm Consortium et al., 2018), as AN correlates negatively with weight parameters on a symptomatic- and genetic level (Duncan et al., 2017; Speranza et al., 2001). OCD and ASD showed a positive correlation with education parameters and OCD correlated negatively with age at first birth (there was no significant correlation between ASD and age at first birth), while the pattern was reversed for ADHD and MDD. Dalsgaard et al. (2020) recently demonstrated that males with OCD achieved significantly higher school grades than individuals without a psychiatric disorder, while people with other psychiatric disorders (except AN) had significantly lower grades. It was also shown that higher education and socio-economic status are associated with higher maternal age at first birth (Van Roode et al., 2017) and that children of young mothers were disadvantaged in schooling (Fall et al., 2015).

Because the four disorders showed differential genetic correlation patterns, we presumed that the polygenic architecture of comorbid OCD subgroups would vary depending on their comorbid diagnosis. We first looked at heritability estimates and genetic correlations between the comorbid OCD subgroups. The *onlyOCD* and the *MC* group demonstrated the highest heritability estimates, while the *OCD+ASD* group displayed the lowest heritability estimates compared to all other subgroups. As sample sizes in each comorbidity group were quite low, SE were generally high and not all of the heritability estimates and none of the genetic correlation estimates between the comorbid subgroups reached significance.

In a last step we then applied PRS analysis across the iPSYCH OCD comorbid subgroups. Rather than selecting traits used as training datasets on a theoretical or clinical background, they were selected in view of their different correlation patterns with OCD, MDD, ADHD, and ASD across a wide range of psychiatric and somatic phenotypes, as we wanted to explore whether the different directions of correlations would be mirrored in the PRS analysis of the OCD comorbid subgroups. For traits for which OCD, MDD, ADHD, and ASD showed a heterogeneous genetic correlation pattern (EA, AFB, BMI) we hypothesized that PRSes based on those traits would also exhibit a heterogeneous pattern of association with the comorbid OCD subgroups. For EA and AFB the pattern of PRS loadings that emerged across the OCD comorbid subgroups closely mirrored the concordance structure of the genetic correlations between OCD and MDD, ADHD, and ASD. OCD and ASD correlated positively with *Years of schooling* and *College or university degree*, while it was the opposite for ADHD and MDD. Accordingly, in the PRS analysis the *OCD+ADHD* group had the highest negative loading for EA, while the EA PRS estimate was positive in the *OCD+ASD* group. Similarities between the correlation analysis and PRS analysis could also be shown for AFB. OCD correlated positively, MDD and ADHD negatively with AFB. ASD did not show a significant correlation. Similarly, in the PRS analysis, AFB was positively associated with disease status in the *onlyOCD* group and to a lower degree also in the *OCD+ASD* group, while it was negatively associated with the *OCD+MDD, OCD+ADHD* and *MC* group. The PRS loadings for BMI was the most negative for the *onlyOCD* group, but also showed a negative association with *OCD+ASD* and *MC*, while, somewhat surprisingly, the other OCD subgroups were not significantly associated with the BMI PRS. One possible explanation for this pattern may be that the negative correlation between OCD and BMI and the positive correlations between ADHD and BMI, as well as between MDD and BMI translate into a null-finding in the PRS finding for BMI because the opposing correlations may evoke counteracting effects in the comorbid subgroups. As neuroticism showed a fairly homogeneous correlation with OCD, MDD, ADHD, and ASD, we expected no polygenic heterogeneity across comorbid OCD subgroups. Similarly, for AN and BP, which correlated significantly positively with OCD and MDD, and positively but non-significantly with ASD and ADHD, we expected a rather homogeneous pattern of association with PRSs across the subgroups, with stronger associations for *onlyOCD* and *OCD+MDD*. This was indeed the case, as PRSes of neuroticism, and BP were associated with OCD across all comorbid subgroups with no significant differences in estimates between the OCD comorbid subgroups. For AN, the pattern of correlations was mirrored closely in the PRS analysis - *onlyOCD* and *OCD+MDD* demonstrated the highest PRS estimates, followed by *OCD+ASD* and *OCD+ADHD*, with a significant difference between the highest estimate for *onlyOCD* and the lowest estimate for *OCD+ADHD*. Because we were also interested how PRS estimates change for traits which showed no correlation with OCD but with some of the other three disorders, we also included insomnia in the PRS analysis. While the insomnia PRS was not significantly associated with the *onlyOCD* subgroup, it showed significant associations with the *OCD+MDD, OCD+ADHD*, and *MC* subgroups, indicating that a comorbid diagnosis might change the association of OCD and insomnia.

Previously, This builds upon results by Hirschtritt et al. (2018) who, using a different approach, examined OCD- and ADHD-symptom dimensions in TS cases and identified unique symptom subgroups that were differentially associated with other comorbid psychiatric disorders.

To conclude, the different PRS estimates across OCD subsets provide the first evidence for a heterogeneous and qualitatively different genetic architecture of OCD subgroups defined by a comorbid diagnosis of MDD, ADHD, and/or ASD. Traits that show a heterogeneous genetic correlation pattern with OCD, MDD, ADHD, and ASD generally also exhibit a heterogeneous pattern of estimations in PRS analysis across OCD comorbid subgroups. This was especially shown for AFB, and EA. While being unique in its approach, results of the present study are in accordance with previous research by Hirschtritt et al. (2018) who examined OCD- and ADHD-symptom dimensions in TS cases and identified unique OCD symptom subgroups that were differentially associated with other comorbid psychiatric disorders. Both, OCD symptom subgroups and comorbid subgroups, may be markers of distinct underlying patterns of psychopathology and genetic architecture.

Because heterogeneous genetic architectures could potentially point towards heterogeneous disease mechanisms, the context in which OCD occurs may have implications for diagnostic criteria and treatment that might not have been considered sufficiently in past and present research and clinical practice. Pallanti et al. (2011), for example, showed that OCD in the presence of comorbid conditions is often associated with non-response to treatment, indicating differential clinical characteristics. Also, for the success of GWAS analyses, it may be beneficial to focus on (sub)phenotype definitions rather than solely relying on increasing sample size. As Kulminski et al. (2016) and MacRae and Vasan (2011) have discussed, increasing the size of many human disease cohorts is likely only to upscale the heterogeneity in parallel. Especially for cross-disorder GWAS analyses, which have gained a lot of attention recently (Cross-Disorder Group of the Psychiatric Genomics Consortium, 2013; Lee et al., 2019; Grotzinger et al., 2019; Abdellaoui et al., 2020), it may be crucial to account for comorbidities to avoid confounding of genetic similarities and differences between psychiatric disorders. One limitation of this study is the right censoring of comorbidities. While ADHD and ASD are neurodevelopmental disorders with mostly a childhood onset (and some persistency into adulthood), MDD usually occurs with an onset in late adolescence and adulthood. Therefore, the possibility that an individual develops a comorbidity, or another comorbidity on top of an already existing one, cannot be ruled out and may be higher for disorders with a later age of onset. Inherently, iPSYCH is a longitudinal study. As with other studies, however, it may be the case that some study participants (e.g. those originally ascertained for their ADHD and/or ASD diagnose) were included at a time point at which the follow-up time was not sufficient to capture a later diagnosis of one of the comorbidities under study (e.g. MDD). While right censoring may dampen some of the observed effects, it is unlikely to alter the overall observations of this study and its main finding of a heterogeneous genetic architecture of comorbid subgroups.

The present study should be viewed as a pilot study and exploratory in nature. In the future, it would be of interest to conduct similar analyses with a broader range of correlated phenotypes and to include other related and comorbid disorders, such as schizophrenia, BP, AN, Tourette’s syndrome and anxiety disorders. It has also been suggested that the onset of OCD (early vs. late) (Hemmings et al., 2004; Walitza et al., 2010; Taylor, 2011), sex (male vs. female) (Khramtsova et al., 2019), or different symptom dimensions of OCD (Hasler et al., 2005) present differing underlying genetic architectures.

## Data Availability

The data used in this study can be made available upon request to the authors (AB).

## 5 CONFLICT OF INTEREST STATEMENT

The authors do not report any conflict of interest.

## 6 AUTHOR CONTRIBUTIONS

NS and MM contributed to the conception and design of the study. AB, OM, MN, TW, DH, and PM contributed to the conception and design of the original iPSYCH study. SM, MBH, JBN, TDA, MH, JC, and JBG contributed to the data collection and organization of the database. NS, JG, and TDA performed the statistical analysis. NS wrote the first draft of the manuscript. All authors contributed to manuscript revision, read, and approved the submitted version.

## 7 FUNDING

The iPSYCH team is funded by the Lundbeck Foundation (R102-A9118, R155-2014-1724, and R248-2017-2003), the EU H2020 Program (Grant No. 667302, “CoCA”), NIMH (1U01MH109514-01) and the universities and university hospitals of Aarhus and Copenhagen.

## 8 ACKNOWLEDGMENTS

This research has been conducted using the Danish National Biobank resource, supported by the Novo Nordisk Foundation. High-performance computer capacity for handling and statistical analysis of iPSYCH data on the GenomeDK HPC facility was provided by the Center for Genomics and Personalized Medicine and the Centre for Integrative Sequencing, iSEQ, Aarhus University, Denmark.

## 9 DATA AVAILABILITY STATEMENT

The data used in this study can be made available upon request to the authors (AB).

## Notes

### Competing Interest Statement

The authors have declared no competing interest.

### Author Declarations

The study was approved by the Regional Scientific Ethics Committee in Denmark and the Danish Data Protection Agency.

