## Supplementary Figures for "Polygenic heterogeneity across obsessive-compulsive disorder subgroups defined by a comorbid diagnosis"

### *Supplementary Material*

#### 1 SUPPLEMENTARY FIGURES

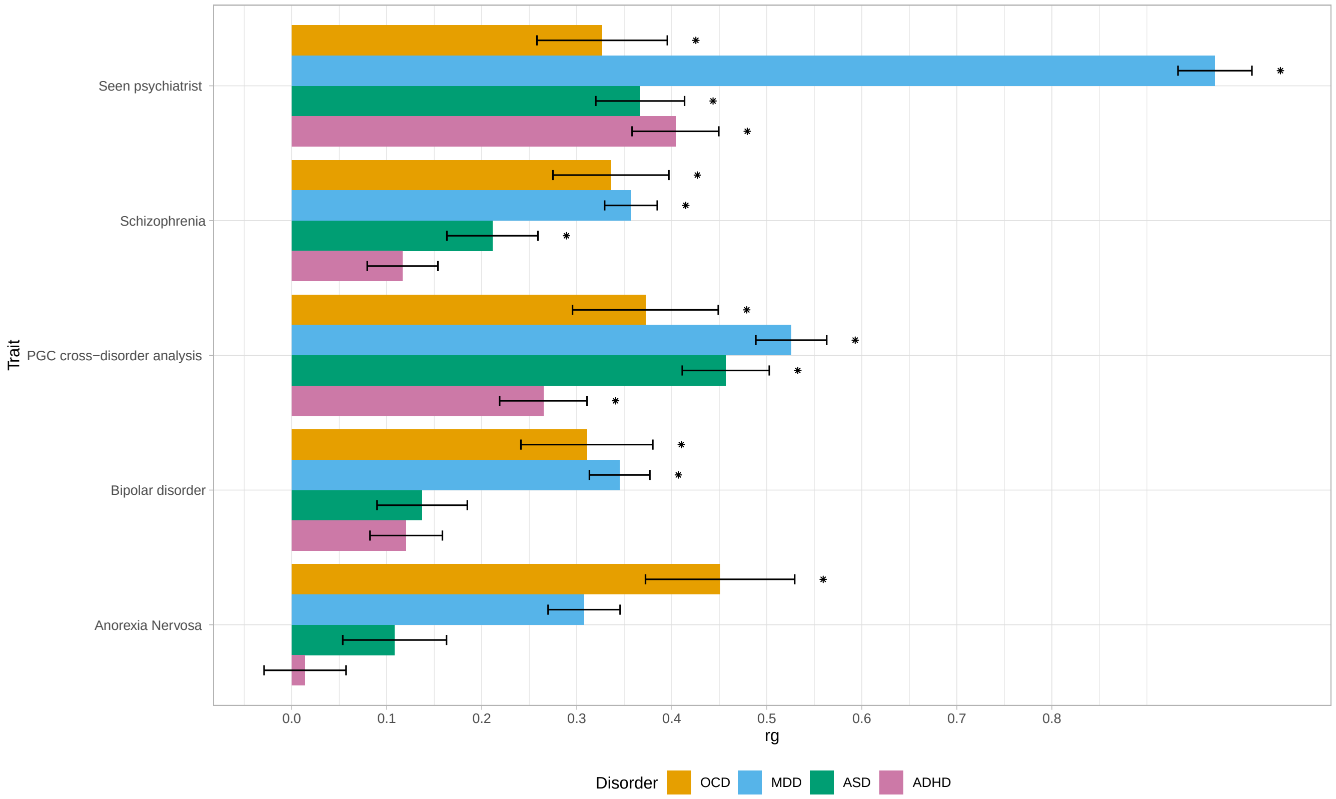


**Figure S1.** Genetic correlation patterns of OCD, MDD, ADHD, and ASD with psychiatric parameters. Bivariate LD score regression was used for the analysis. * indicates statistical significance at a Bonferroni-corrected threshold of 6.42x$10^{-5}$ corrected for 779 tests. The phenotypes correspond to the following data sets: seen a psychiatrist for nerves, anxiety, tension or depression (Neale, 2018), schizophrenia (Ripke et al., 2014), PGC cross-disorder analysis (Cross-Disorder Group of the Psychiatric Genomics Consortium, 2013), bipolar disorder (Stahl et al., 2019), and anorexia nervosa (Watson et al., 2019).

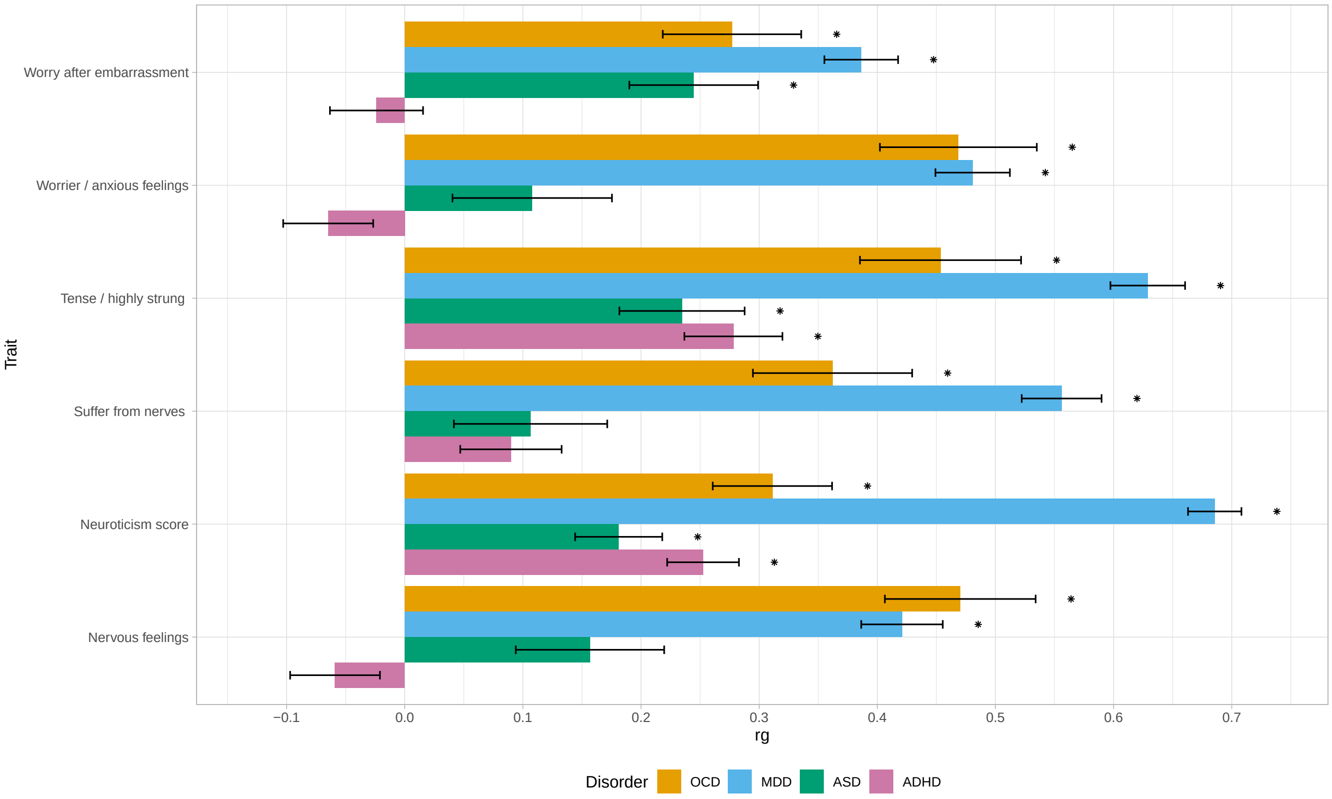


**Figure S2.** Genetic correlation patterns of OCD, MDD, ADHD, and ASD with personality and psychological parameters. Bivariate LD score regression was used for the analysis. * indicates statistical significance at a Bonferroni-corrected threshold of $6.42x10^{-5}$ corrected for 779 tests. The phenotypes correspond to the following data sets: worry after embarrassment (Neale, 2018), worrier/anxious feelings (Neale, 2018), tense/highly strung (Neale, 2018), suffer from nerves (Neale, 2018), neuroticism score (Nagel et al., 2018), and nervous feelings (Neale, 2018).

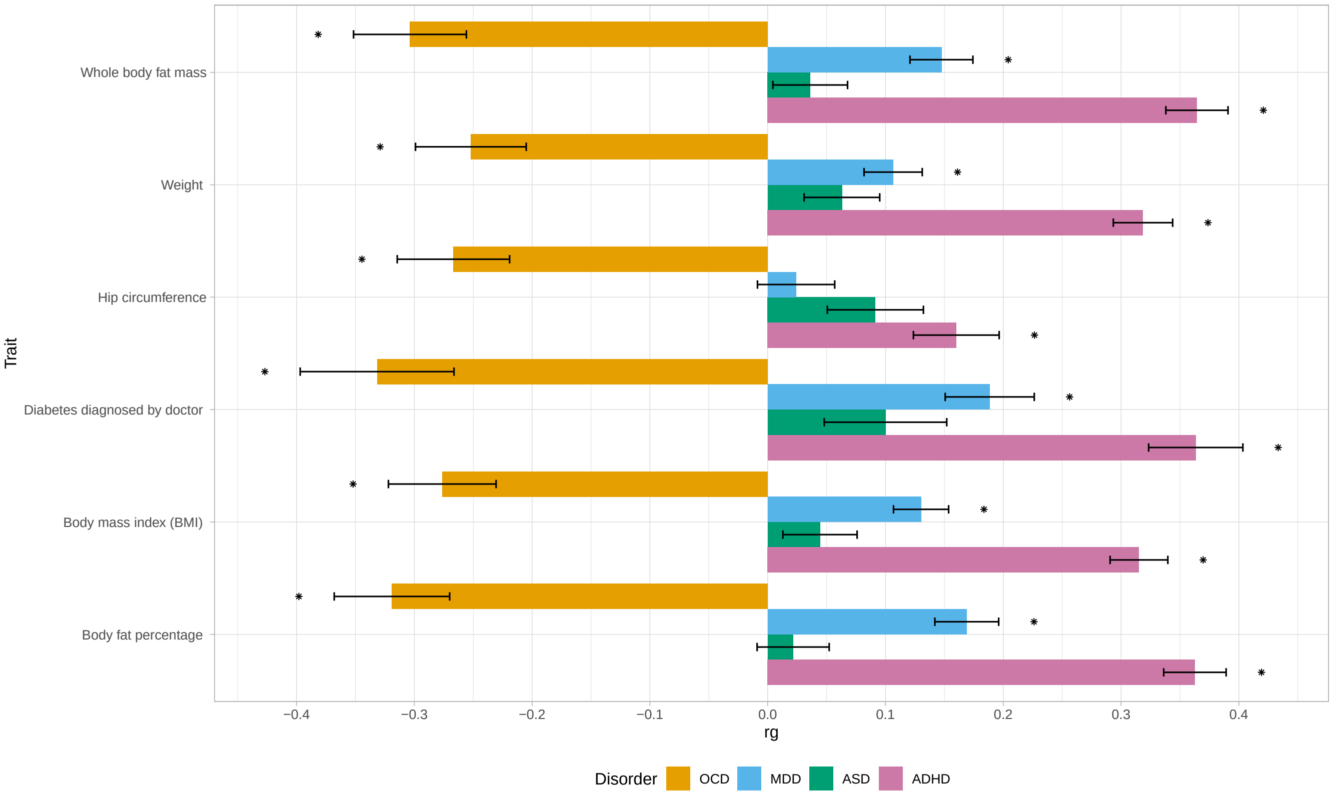


**Figure S3.** Genetic correlation patterns of OCD, MDD, ADHD, and ASD with anthropomorphic and metabolic parameters. Bivariate LD score regression was used for the analysis. * indicates statistical significance at a Bonferroni-corrected threshold of $6.42 \times 10^{-5}$ corrected for 779 testss. The phenotypes correspond to the following data sets: Whole body fat (Neale, 2018), weight (Neale, 2018), hip circumference (Neale, 2018), diabetes diagnosed by doctor (Neale, 2018), body mass index (BMI) (Yengo et al., 2018), and body fat percentage (Neale, 2018).

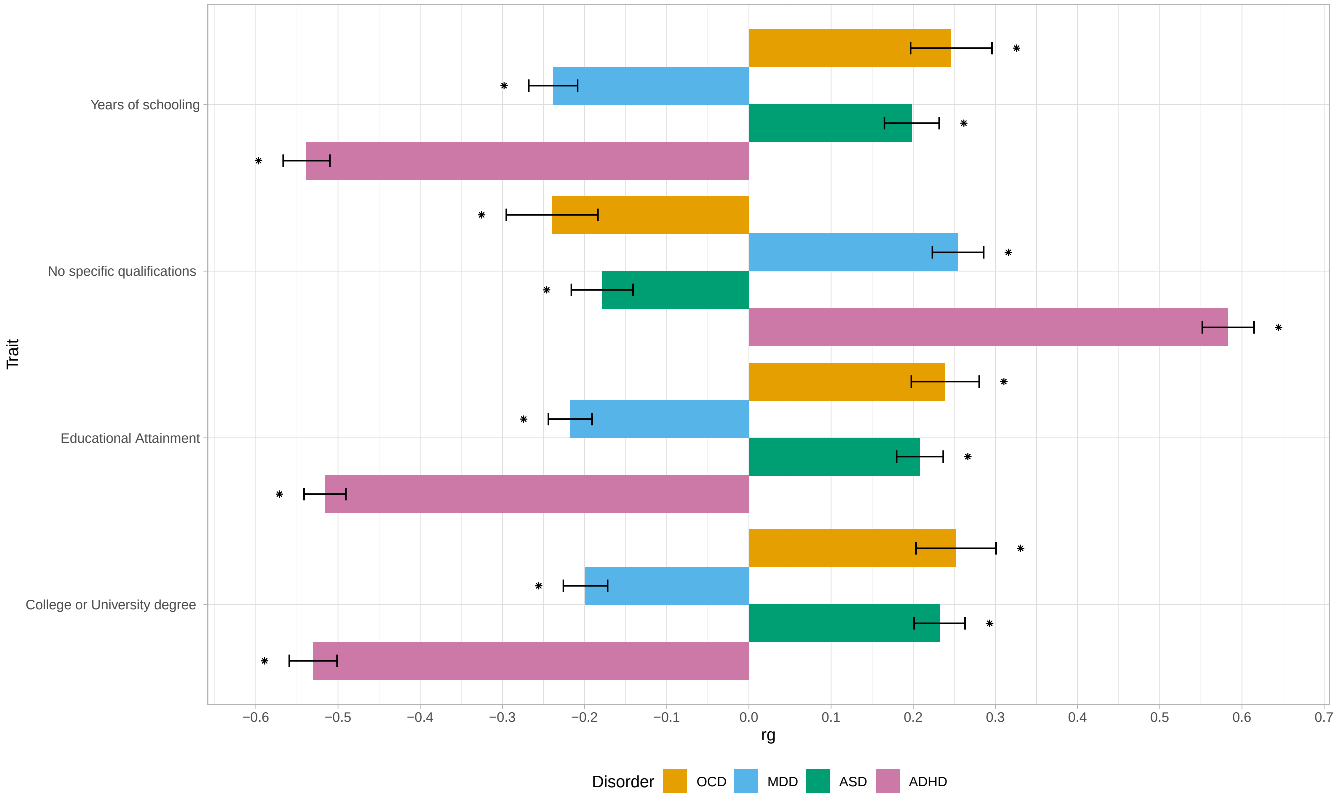


**Figure S4.** Genetic correlation patterns of OCD, MDD, ADHD, and ASD with education parameters. Bivariate LD score regression was used for the analysis. * indicates statistical significance at a Bonferroni-corrected threshold of 6.42x$10^{-5}$ corrected for 779 tests. The phenotypes correspond to the following data sets: years of schooling (Okbay et al., 2016), no specific qualifications (Neale, 2018), educational attainment (Lee et al., 2018), college or university degree (Neale, 2018).

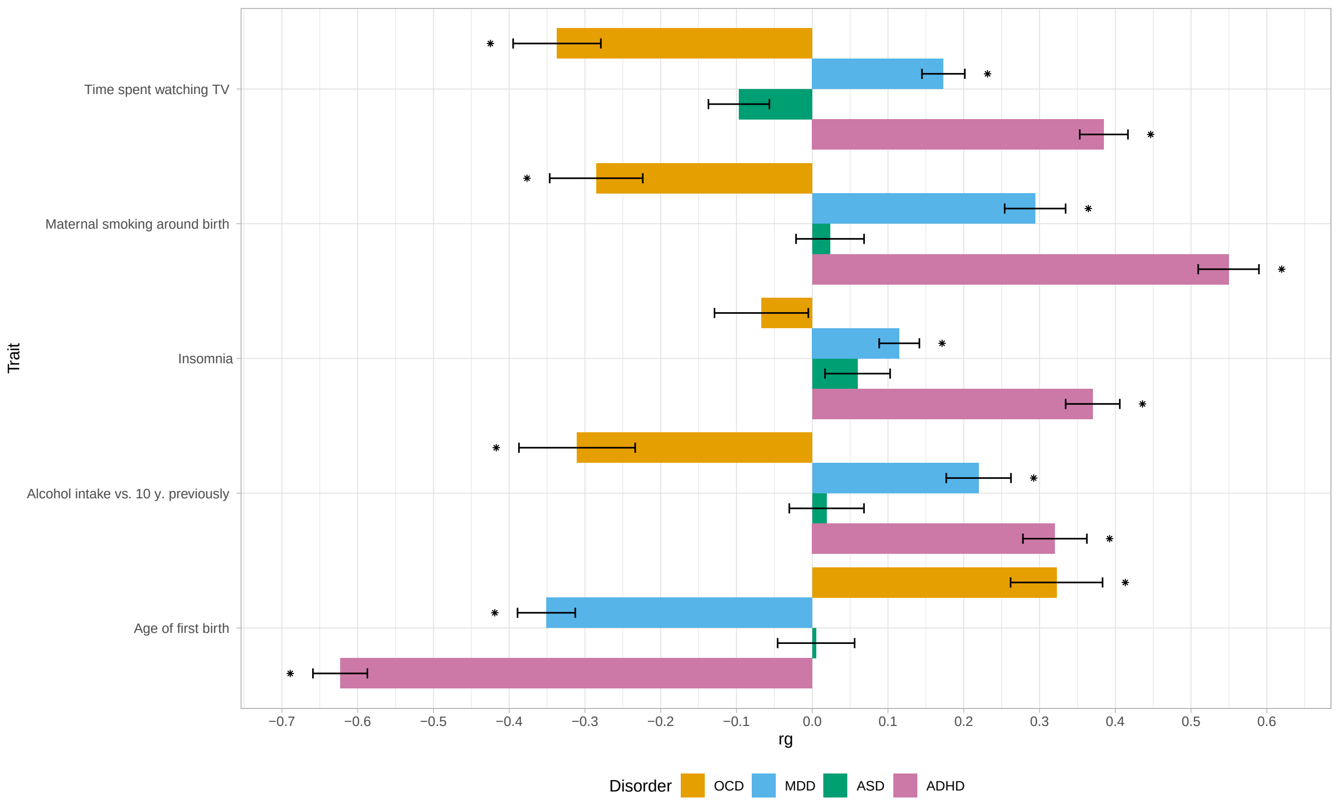


**Figure S5.** Genetic correlation patterns of OCD, MDD, ADHD, and ASD with other health risk parameters. Bivariate LD score regression was used for the analysis. * indicates statistical significance at a Bonferroni-corrected threshold of 6.42x$10^{-5}$ corrected for 779 tests. The phenotypes correspond to the following data sets: time spent watching TV (Neale, 2018), maternal smoking after birth (Neale, 2018), insomnia (Jansen et al., 2019), alcohol intake vs. 10 years previously (Neale, 2018), and age at first birth (Barban et al., 2016).
